# Peripheral metabolic signatures in patients with neuropsychiatric long COVID syndrome

**DOI:** 10.64898/2026.08.07.26359942

**Authors:** Dario Lucas Bergmann, Sophie Neugebauer, Tonia Rocktäschel, Eva-Maria Dommaschk, Meng Li, Alexander Weuthen, Alexander Refisch, Nikolai Blekic, Michael Kiehntopf, André Scherag, Helgi B. Schiöth, Chai K. Lim, Nils Opel, Martin Walter, Bianca Besteher

**Author notes:** <u>Correspondence should go to:</u> PD Dr. Bianca Besteher, Department of Psychiatry and Psychotherapy, Jena University Hospital, Friedrich Schiller University Jena, Philosophenweg 3, 07743 Jena, Germany. Email addresses of all authors: Dario Lucas Bergmann, Sophie Neugebauer, Tonia Rocktäschel, Eva-Maria Dommaschk, Meng Li, Alexander Weuthen, Alexander Refisch, Nikolai Blekic, Michael Kiehntopf, André Scherag, Helgi Schiöth, Chai K. Lim, Nils Opel, Martin Walter.

## Abstract

Neuropsychiatric symptoms are considered the most common feature of long COVID disease. Recent studies have demonstrated structural brain changes and highlighted the importance of neuroinflammation in the development of cognitive deficits as seen in long COVID patients. In addition, peripheral studies have demonstrated heterogeneous molecular subtypes of long COVID pathology. However, it is unknown which peripheral metabolomic alterations occur in patients with neuropsychiatric long COVID syndrome and how these relate to symptom severity. In the present study, we investigated differences in the peripheral serum metabolome profiles of healthy controls and patients with long COVID syndrome with neuropsychiatric symptoms. We found that patients with long COVID showed peripheral alterations in lipid species such as triacylglycerides and acylcarnitines. Furthermore, metabolites altered in patients with long COVID syndrome were also associated with depressive and fatigue symptom burden as well as with differences in cortical thickness in multiple brain regions. Our results demonstrate a metabolic phenotype of long COVID patients that may reflect a dysregulation of lipid metabolism and deficits in mitochondrial energy production as potential contributors to symptom burden and brain structural alterations. These data may serve as a resource and basis for further studies aimed at investigating peripheral molecular alterations in patients with neuropsychiatric long COVID syndrome.

## I. Introduction

Post-acute sequelae of infections with the SARS-CoV-2 virus (PASC) that persist for more than 4 weeks are subsumed under the term “long COVID syndrome” (1). Symptoms can manifest in nearly all organs (2), however, neuropsychiatric symptoms such as fatigue, cognitive impairment (e.g. concentrations and memory deficits) and depressed mood belong to the most common symptoms that evoke severe mental suffering for affected individuals (3). According to Iwasaki et al., long COVID syndrome is clinically almost indistinguishable from other so called post-acute infection syndromes (PAIS) that occur after other viral infections such as after influenza A virus and varicella zoster virus infections, which share fatigue, cognitive deficits and depressed mood as leading symptoms (1, 4, 5). If clinical symptoms persist and the Canadian consensus criteria are fulfilled (6), patients with long COVID syndrome can also be diagnosed with myalgic encephalomyelitis/chronic fatigue syndrome (ME/ CFS) as both diseases may share a similar pathophysiological basis characterised by peripheral endocrinological, metabolic and immunological changes, as reviewed elsewhere (7). Patients with ME/CFS systemically display an impairment in ATP production accompanied by a “hypometabolic state” and an imbalance of pro-and antioxidant metabolites towards more pro-oxidant processes (8, 9). This hypometabolic state is characterized by reduced oxidation of fatty acids via the TCA cycle (8, 9), which has already been observed already in long COVID patients during exercise (10). Acylcarnitines have been identified as significant regulators of fatty acid oxidation. These metabolites facilitate the import of acyl groups derived from fatty acids into mitochondria for subsequent beta-oxidation and are important modulators of different CNS processes (11). Short-chain acylcarnitines have been hypothesised to exert a neuroprotective effect by supporting neuronal function, while medium- and long-chain acylcarnitines are believed to be rather harmful (11). Interestingly, in ME/CFS patients a decrease in short-chain acylcarnitines and an increase in medium-and long-chain acylcarnitines has been demonstrated recently (12).

Furthermore, several -omics studies of long COVID syndrome have been published and reported in part conflicting data in terms of in-/decreases in acylcarnitine species of different length (13, 14). To our knowledge the largest plasma metabolomics study conducted so far in long COVID patients focussed in their results primarily on a decrease in cortisol 2-3 months post-infection (15). However, further results can be observed in their dataset, such as a downregulation of acetylcarnitine, as we have demonstrated by re-analyzing the Su et al. dataset in previous work (16), which may be an indicator of decreased mitochondrial energy production. A more recent study also investigated metabolomic and proteomic profiles of neuropsychiatric long COVID patients in the cerebrospinal fluid, exposing also central metabolomic changes, primarily a disruption in sphingolipid metabolism (17). However, to the best of our knowledge, a systematic analysis of peripheral metabolomic profiles in a patient cohort with neuropsychiatric symptoms, and an analysis of the molecular and metabolomic correlates of neuropsychiatric symptoms, has not yet been conducted. Therefore, we aimed to investigate systemic alterations in peripheral serum metabolome profiles in patients with neuropsychiatric long COVID syndrome (“PC”) compared to healthy survivors of SARS-CoV-2 virus infection (“HC_S”) and healthy never-infected controls (“HC_N”). To this end, we used targeted mass spectrometry-based metabolomics to measure between-group differences in serum concentrations of ∼630 metabolite and lipid molecules. Furthermore, we aimed to investigate potential associations of those metabolites that were altered between the three experimental cohorts with clinical symptom severity of depressive mood, cognitive deficits and memory deficits. Finally, we wanted to investigate potential associations of those metabolites that were altered between cohorts with differences in cortical thickness in multple brain regions.

## II. Materials and Methods

### Participants and cohort phenotyping

Patients were recruited from the post-COVID outpatient clinic of the Department of Internal Medicine IV (Infectiology) and the Department of Neurology at Jena University Hospital. There, they underwent verification of their post-COVID condition via real-time reverse transcriptase-polymerase chain reaction (RT-qPCR) at the time of acute infection, as well as a medical history assessment conducted by a board-certified physician, covering the timepoint and severity of their COVID-19 symptoms according to the WHO criteria. The long COVID condition was defined according to the S1 guideline as the presence of persistent symptoms 4 to 12 weeks after the initial SARS-CoV-2 infection, while post-COVID syndrome was classified as the persistence of symptoms beyond 12 weeks (18). In the cohort described in this study, the average time since the initial SARS-CoV-2 infection was 10.5 months (range: 3.5 – 24.5 months). Inclusion criteria did not impose restrictions regarding specific symptoms or recovery time to ensure coverage of the full spectrum of long-COVID symptomatology. None of the participants met the criteria for any DSM-5 disorder at the time of assessment, as determined through careful screening using the MINI Interview, conducted by a trained rater according to DSM-5 criteria (19). Additionally, participants had no history of major neurological or untreated medical conditions. Exclusion criteria for all participants included: general MRI contraindications, neurological conditions, and untreated internal medical conditions, particularly chronic inflammatory diseases, as well as a history of/or current substance use disorder. All participants completed the Multiple-Choice Vocabulary Test B (MWT-B), which is available in German (20), to estimate crystallized IQ and confirm the inclusion criterion of an IQ above 80. All participants provided written informed consent before participating in the study. The study protocol was approved by the local Ethics Committee of Jena University Medical School. Between April 2021 and June 2022, we included 54 long-COVID patients, 18 healthy survivors who had laboratory-confirmed COVD-19 but did not develop long-COVID syndrome, and 30 healthy controls without known or serologically confirmed prior COVD-19 infection from whom we measured metabolomic profiles. The groups were matched for age and sex. A summary of demographic characteristics of groups is given in Table 1.

**Table 1.**
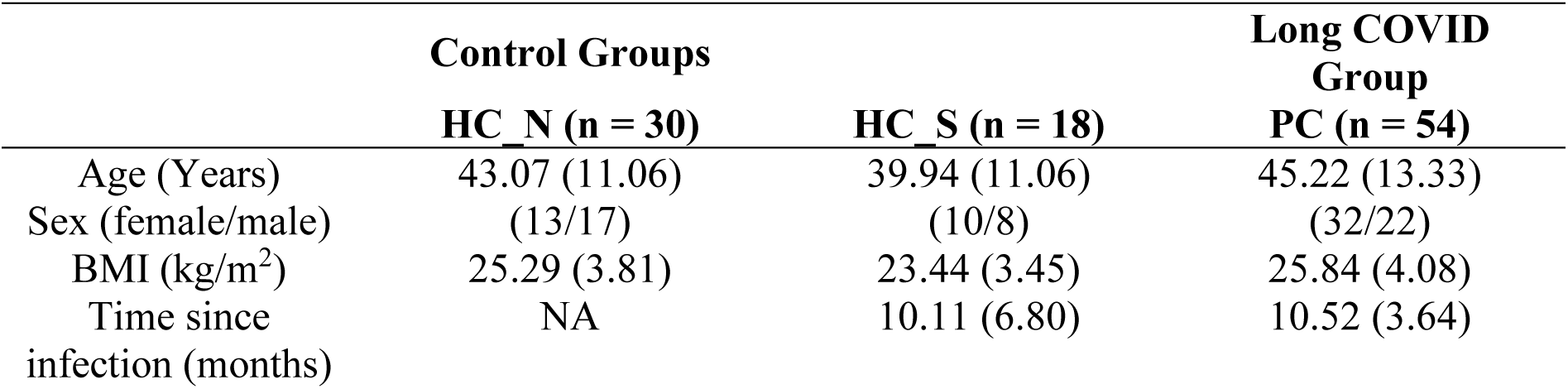
Subject demographic data. Shown are arithmetic mean and standard deviation in brackets.

For the assessment of specific symptom scores, established psychological tests available in German were utilized. The Short Form-36 Health Survey (SF-36), a self-report questionnaire, was used to measure general health-related quality of life across eight dimensions: Physical Functioning (PF), Role Limitations due to Physical Health (Role-Physical, RP), Bodily Pain (BP), General Health Perceptions (GH), Vitality/Energy (VIT), Social Functioning (SF), Role Limitations due to Emotional Problems (Role-Emotional, RE), and General Mental Health (MH) (21). Additionally, two composite scores can be derived from these dimensions: the Physical Component Summary (PCS) and the Mental Component Summary (MCS). For neurocognitive screening, the Montreal Cognitive Assessment (MoCA) was administered. A score below 26 indicates mild to moderate cognitive impairment, whereas a score of 26 or higher suggests no cognitive impairment (22, 23). To assess depressive symptoms in the context of post-COVID syndrome, the Beck Depression Inventory-II (BDI-II) was used (24). This self-report questionnaire consists of 21 items, each rated on a 0–3 scale. Scores are interpreted as follows: 0–8 indicates no depressive symptoms, 9–13 minimal symptoms, 14–19 mild depression, 20–28 moderate depression, and 29–63 severe depression. Additionally, depressive symptoms were assessed using the Montgomery-Åsberg Depression Rating Scale (MADRS) (25). This clinician-rated scale evaluates 10 core depressive symptoms, each rated on a 0–6 scale. A total score of 0–6 indicates no depression, 7–19 mild depressive symptoms, 20–34 moderate depression, and above 34 severe depression.

### Magnetic resonance imaging (MRI) and calculation of cortical thickness

MRI data obtained in this study were published before (26). Subjects underwent high–resolution T1-weighted MRI on a 3 Tesla Siemens Prisma fit scanner (Siemens, Erlangen, Germany) using a standard quadrature head coil and an axial 3-dimensional magnetization. prepared rapid gradient echo (MP-RAGE) sequence (TR 2400 ms, TE 2.22 ms, α 8◦, 208 contiguous sagittal slices, FoV 256 mm, voxel resolution 0.8 × 0.8 × 0.8 mm; acquisition time 6:38 min). All subjects gave their consent for the native brain MRI scan after consultation of a licensed radiologist. The scan was part of an MRI protocol of 60 min total duration. All scans were checked to exclude imaging artefacts. Results reported in this manuscript derive from preprocessing performed using fMRIPrep 23.2.1(27). FreeSurfer’s recon-all pipeline (FreeSurfer 7.3.2)(28, 29), integrated within fMRIPrep, was used for cortical reconstruction and segmentation. This pipeline generates cortical parcellations based on the Desikan-Killiany atlas (commonly referred to as aparc or DK40)(30), typically partitioning the cortex into 68 regions across both hemispheres. Average cortical thickness values for each region were then extracted using FreeSurfer’s aparcstats2table command, yielding robust morphometric metrics for subsequent analysis. The “ggseg” R package (31) was used to visualise the regression analysis estimates of cortical thickness across the DK40 region in relation to specific metabolites.

### Blood collection and preanalytics

Standardized blood collection was performed on all participants within a designated time frame between 7:30 and 9:00 a.m. on the day of MRI scanning. Serum gel monovettes (7.5 mL, Sarstedt) were used, and on average 30 minutes after collection (range 20-40 minutes), samples were centrifuged at 800 g for 10 minutes at 4°C. The supernatant was then divided into aliquots of 0.25 mL each and stored at −80°C. Frozen samples were subsequently transferred to the Institute of Clinical Chemistry for further analysis, without any further freeze-thaw cycles.

### Targeted metabolomics – sample preparation and mass spectrometry

The concentrations of up to 630 metabolites from 26 biochemical classes (40 acylcarnitines [AC], 1 amine oxide [AO], 1 alkaloid [Al], 20 amino Acids [AA], 30 amino acid related [AR], 14 bile acids [BL], 9 biogenic amines [BA], 7 carboxylic acids [CA], 1 carbohydrate and related [CR], 28 ceramides [Cer], 22 cholesteryl esters [CE], 1 cresol [CL], 44 diglycerides [DG], 9 dihexosylceramides [DH], 8 dihydroceramides [DC], 12 fatty acids [FA], 19 hexosylceramides [HC], 4 hormones [H], 4 indoles and derivates [ID], 14 lysophosphatidylcholines [LP], 2 nucleobases and related [NR], 76 phosphatidylcholines [PC], 15 sphingomyelins [SM], 242 triglycerides [TG], 6 trihexosylceramides [TH], and 1 vitamin and cofactor [VC] were quantified in serum following the manufacturer’s protocol (MxP® Quant 500 kit, Biocrates Life Science AG, Innsbruck, Austria). Analyses were performed using a QTrap 5500 LC-MS/MS-System (AB Sciex, Framingham, MA, USA) equipped with an Nexera UHPLC-System (Shimadzu, Japan) and operated with Analyst 1.7.3 software (AB Sciex, Framingham, MA, USA). For sample preparation, 10 µl of calibration standard, quality control, zero sample, or study sample were applied to the center of the spots on the kit plate and dried under nitrogen for 30 minutes at room temperature. Subsequently, 50 µl of a 5% phenylisothiocyanate solution (Merck, Darmstadt, Germany) was added, incubated for 1 hour at room temperature, and dried again for 1 hour using the nitrogen evaporator. Metabolites were then extracted with 300 µl of a 5 mM ammonium acetate solution in methanol (Merck, Darmstadt, Germany; Roth, Karlsruhe, Germany) and incubated for 30 minutes on a shaker at 450 rpm. The plate was centrifuged at 100 g for 2 minutes, and the upper plate was removed. The resulting extracts were divided for two separate analytical approaches: 150 µl for LC-MS and 10 µl for flow injection analysis (FIA). An additional 150 µl of HPLC-grade water was added to the LC-MS plate, while 490 µl of FIA solvent (Biocrates solvent diluted in methanol) was added to the FIA plate. The LC-MS analysis was prioritized using multiple reaction monitoring (MRM), while the FIA plate was stored at 4°C. For LC-MS, 5 µl of each sample was injected twice (once for positive and once for negative ionization mode) onto the kit column and eluted using solvent A (HPLC water + 0.2% formic acid) and solvent B (acetonitrile + 0.2% formic acid). FIA analysis involved two successive 20 µl injections directly into the MS at a flow rate of 30 µl/min using FIA solvent. Metabolite concentrations were determined using the MetIDQ™ software package (integrated with the MxP® Quant 500 kit) by comparing analyte/internal standard ratios or measured analytes in a defined extracted ion count section to those of the calibration curve or specific labeled internal standards or nonlabelled, nonphysiologically standards (semiquantitative) provided with the kit. Data were then normalized sample-wise using quality controls and exported for further analysis.

### Metabolomics bioinformatic analysis

#### Data preprocessing

The obtained metabolite concentrations in µmol/l were further processed by filtering for all metabolites that had ≥ 80% missing values in all cohorts, to reduce the effect of missing values, which led to the removal of 6 out of 630 metabolites. Subsequently, concentrations were log_2_-transformed. As the reason for missing values was that in the respective sample, the value was below the detection limit and thus the data are “missing not at random” (MNAR) (32), we used quantile regression imputation of left-censored data (QRILC), as this method performs best for left-censored MNAR data (33). QRILC was done using the impute.QRILC function from the “imputeLCMD”-package in R (34), with a sigma value of 0.5 as most of the data did not follow a gaussian normal distribution, as determined by Shapiro-Wilk tests, as suggested previously (32). The annotation of metabolite names to HMDB IDs, full names and class, was performed using the keytables provided by *Biocrates* with the MxP® Quant 500 kit.

### Differential abundance, pathway enrichment analysis and data visualization

In order to identify differentially abundant metabolites (DAMs), we used the lme4 R package (35) and therein linear mixed models that included age, sex and BMI and all interactions of these as covariates. In addition, batch was included as a random effect. Metabolite values were log_2_ transformed, as described previously (15). Subsequently, the multcomp R package (36) was used to determine between cohort differences and fold changes. Specifically, the glht function was applied to calculate p-values for between cohort comparisons, all obtained p-values were false discovery rate adjusted (= q-values). All metabolites that had a p_adj(fdr)_ ≤ 0.1 were used for further analyses. Pathway enrichment was performed using the enrichHMDB function from the MicrobiomeProfiler R package, which is part of the clusterProfiler R package (37). The total metabolome was used as background and all in one of the three between-cohort comparisons changed metabolites (p_adj(fdr)_ ≤ 0.1) were used for enrichment analysis. The obtained, unadjusted, p-values of the top 15 enriched pathways were visualized using a bar plot. Volcano plots and violin plots were generated with customized functions using the ggplot2 package within the tidyverse package. Heatmaps were generated from the z-score transformed metabolite concentrations after covariate adjustment with the ComplexHeatmap R package (38). Agglomerative clustering by average linkage method and Euclidean distance was performed on the rows (i.e. metabolites) and the dendrograms were thereafter reordered using the dendsort R package (39).

### Integration with neuropsychological/psychometric scores and cortical thickness values and validation in the INCOV cohort

All neuropsychological/psychometric scores obtained as well as all cortical thickness values determined as described above, were used to fit linear mixed models as described above, but for each score two models were fitted, one with the interaction between the score and cohort (e.g. “BDI_2*cohort”) in addition to all covariates, and one without the scores. Likelihood ratio tests were then performed to compare the two models for each score and to identify models where the inclusion of the score improved the model fit. All obtained p-values (Pr(>Chisq)) were further false discovery rate (FDR) adjusted and we used p_adj(fdr)_ ≤ 0.1 as a pragmatic threshold for prioritizing candidate metabolite associations with the respective score/cortical thickness of the respective region. Model summaries and estimates for each model variable were obtained using the jtools R package (40). To validate our findings, we used the previously published metabolomic data of PASC patients from the INCOV cohort (15), based on the datasets provided in their publication (Table S2, sheet “S2.2 Metabolomics” and Table S1, sheet “S1.3 PASC data”), as described in the Results section.

### Statistical procedures

All analyses were performed using R 4.5.2 (41) or Graphpad Prism 8.4 (**Figure S1**). Shapiro-Wilk normality test was performed before further statistical analysis was performed. Depending on the result, parametric or non-parametric statistical tests were conducted, as stated in the respective figure legends. Statistical tests for metabolomic analysis and functional annotation and enrichment analyses were performed as indicated in the respective material and methods section. As indicated in the respective methods section or figure legend, p-values obtained from multiple significance tests were appropriately adjusted using false discovery rate (FDR). Statistical significance was accepted if the p_adj_-value (p) was p ≤ 0.1, unless otherwise stated in the respective figure legends.

## III. Results

### Neuropsychiatric long COVID patients display subtle peripheral alterations in lipid metabolism

Our study cohort consisted of 54 patients with long COVID symptomatology, with persisting symptoms of at least 12 weeks post-infection (“PC”), as well as two healthy control groups consisting of adults who had a SARS-CoV-2 infection, but did not develop any long COVID symptoms (“HC_S”) and adults who never had any SARS-CoV-2 infection (“HC_N”) (**Table 1**). The cohorts did not differ significantly in terms of age, sex and BMI, as previously described (26). In addition to blood analysis, the patients included in this study also underwent neuropsychological/psychometric phenotyping, including SF-36 score for assessment of general health and specifically assessment of vitality, as well as MoCA to detect potential cognitive impairments and to assess depressive symptoms BDI-II and MADRS scores were obtained (**Supplemental Dataset S1**). The long COVID cohort analyzed in this study showed on average mild depressive symptoms (Given are arithmetic means, standard deviation in brackets: X̄_BDI-II (PC)_ = 15.5 (8.540), X̄_BDI-II (HC_N)_ = 3.172 (3.685), X̄_BDI-II (HC_S)_ = 7.944 (10.410) and X̄_MADRS (PC)_ = 12.76 (8.221), X̄_MADRS (HC_N)_ = 2.433 (2.300), X̄_MADRS (HC_S)_ = 4.118 (6.480)), a subtle cognitive impairment (X̄_MOCA (PC)_ = 25.93 (2.746), X̄_MOCA (HC_N)_ = 27.83 (2.036), X̄_MOCA (HC_S)_ = 27.65 (2.262)) and a profound decrease in overall quality of life and specifically vitality as a measure for fatigue (X̄_SF_36_GH (PC)_ = 43.33, X̄_SF_36_GH (HC_N)_ = 72.6, X̄_SF_36_GH (HC_S)_ = 72.35 and X̄_SF_36_VIT (PC)_ = 33.21 (20.55), X̄_SF_36_VIT (HC_N)_ = 64.00 (18.21), X̄_SF_36_VIT (HC_S)_ = 61.76 (20.31)), with no differences between the two control groups “HC_N” and “HC_S” (**Figure S1**).

**Figure S1.**
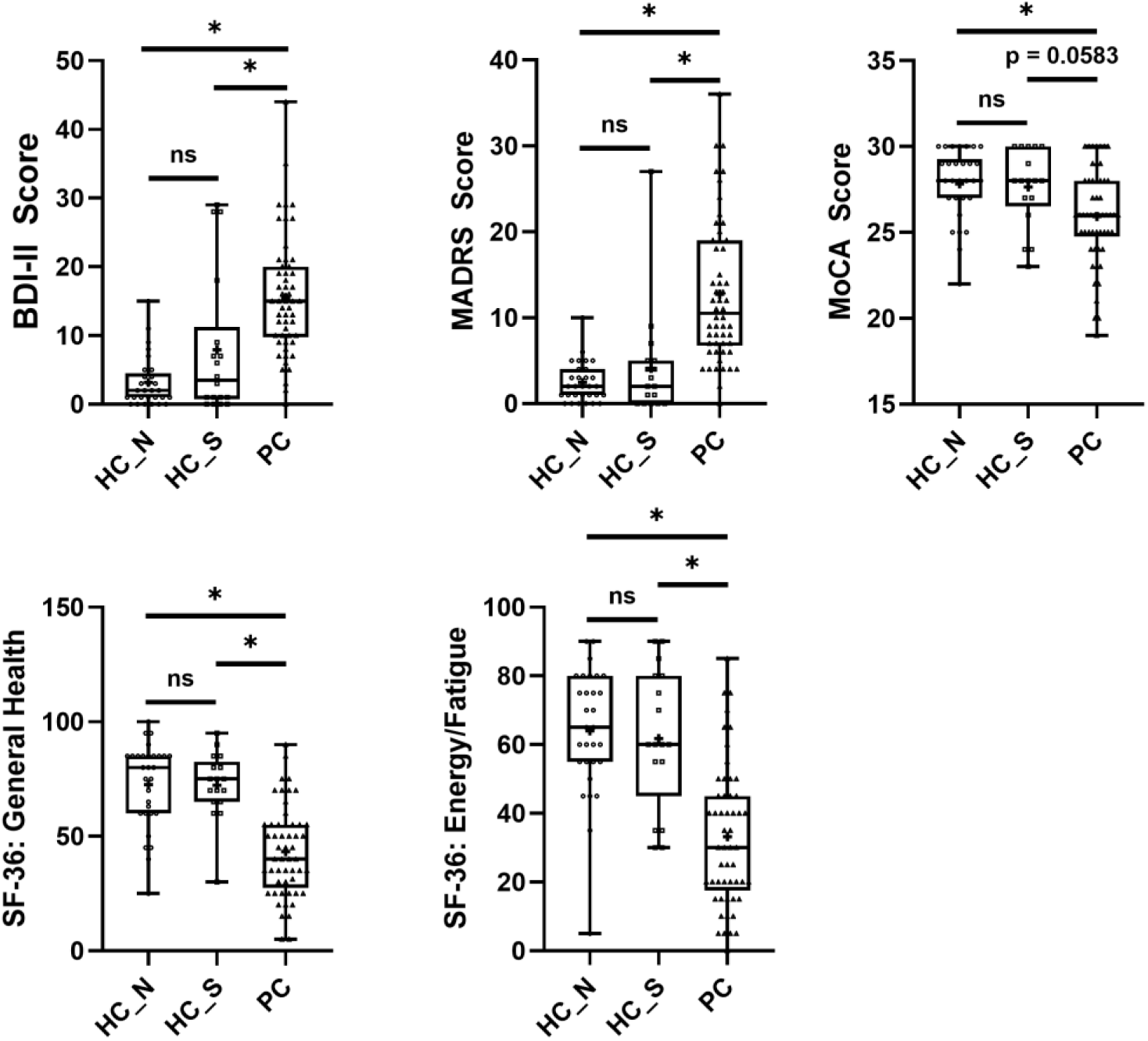
Changes in neuropsychological/psychometric symptom scores in long COVID patients. Boxplots depicting the distribution of individual participant scores for depressive symptoms (BDI-2 and MADRS), cognitive deficits (MoCA) and general health as well as fatigue symptoms (SF-36 GH and VIT). Whiskers represent minimum to maximum distribution, the box shows the interquartile range as well as the median, a “+” depicts the mean. For all comparisons a Kruskal-Wallis test was conducted with a p-value ≤ 0.05 in all conducted tests and with Post-hoc Dunn’s multiple comparisons tests with * = p_adj_ ≤ 0.05 in the respective comparison.

We used linear mixed models to adjust the serum metabolite concentrations for age, sex and BMI and their interactions as critical determinants of inter-individual metabolite variation, as well as batch, as described in other recent metabolomics studies in PASC patients (e.g. (15)) (**Supplemental Dataset S2**). Accordingly, a principal component analysis (PCA) showed no major differences along the first two principal components between sexes (**Figure S2A**), age (**Figure S2B**) and between individuals with low vs. high-BMI after covariate adjustment (**Figure S2C**).

**Figure S2.**
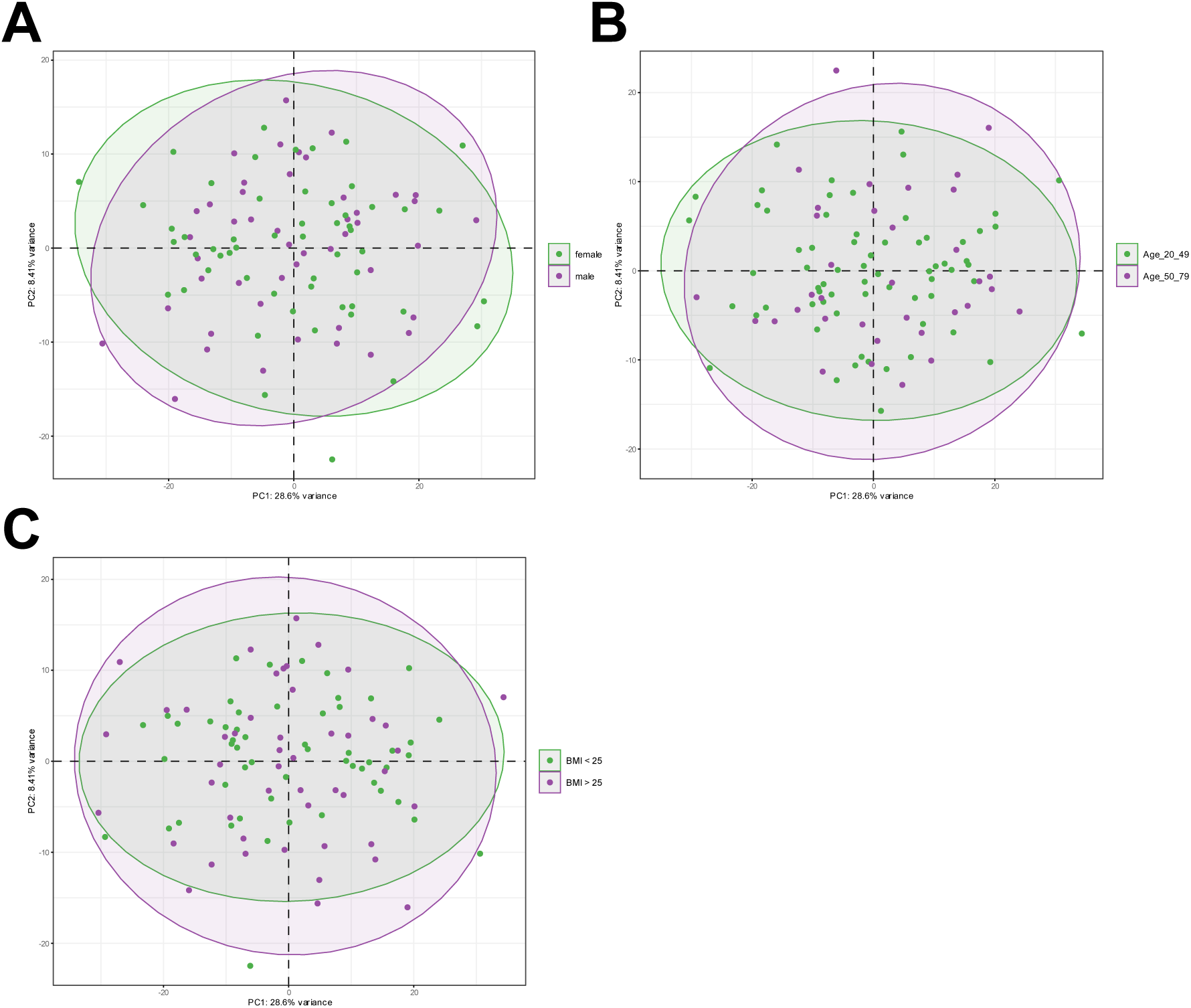
Principal component analysis of the total metabolome after adjustment for age, sex and BMI. **A to C** Principal component analysis from the total metabolome dataset, after adjusting for batch, age, sex and BMI. All individuals from all cohorts are depicted and individuals are coloured according to either their sex (male vs. female, **A**), their age (ranging from either 20 to 49 vs. 50 to 79, **B**) or their BMI (BMI < 25 vs. BMI > 25, **C**). The first principal component axes are depicted for all plots.

However, principal component analysis also revealed no major shifts between the three different cohorts along the first four principal components, accounting for ∼45% of the total metabolome variance), indicating rather subtle differences in the total serum metabolome in long COVID patients compared to healthy controls (**Figure 1A** and **1B**). In addition, we performed hierarchical agglomerative clustering on the z-score transformed and age-, sex- and BMI-adjusted metabolite concentrations as well as within-cohort clustering of samples to identify potential metabolite clusters that differed between cohorts. Indeed, three major clusters are apparent, one cluster that showed no variation within and between cohorts, and two clusters that showed both, within-cohort variation in different types of lipid species as well as subtle changes between cohorts (**Figure 1C**).

**Figure 1.**
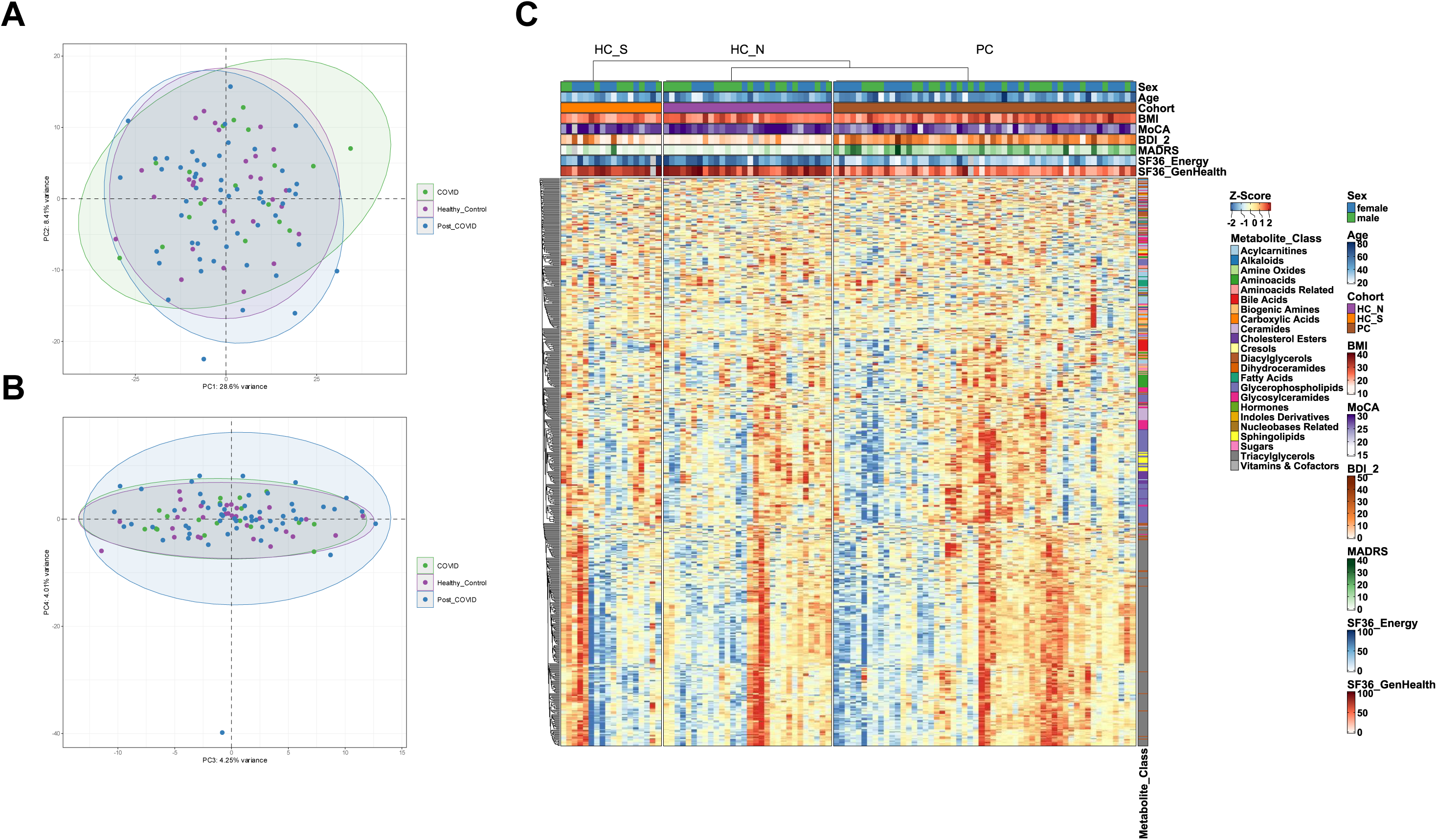
Principal component analysis of the total metabolome and heatmap with hierarchical clustering of metabolites and study participants. **A, B** Principal component analysis from the total metabolome dataset, after adjusting for batch, age, sex and BMI. All individuals from all cohorts are depicted and individuals are coloured according to the cohort (“COVID” = “HC_S”, “Healthy_Control” = “HC_N”, “Post_COVID” = “PC”). The first principal component axes are depicted in **A** and the 3^rd^ and 4^th^ principal component axes are shown in **B**. **C** Visualization of the z-score normalized metabolite concentrations of all measured metabolites of all samples after adjusting for batch, age, sex and BMI. Metabolites were clustered by agglomerative hierarchical clustering with average linkage. In addition, within each cohort, clustering was performed by default with agglomerative hierarchical clustering with complete linkage. In addition, several column and row annotations are shown in the heatmap: The columns are divided by cohort, the covariates age, sex and BMI are shown on different colour scales. Furthermore, the clinical symptom scores (BDI-II, MADRS, MoCA, SF36_Vitality and SF36_General) are annotated on the columns above each heatmap. Lastly, the metabolite class of each metabolite is annotated to the right of the heatmaps.

Having established overarching metabolic differences between cohorts, we next performed a complementary metabolite-level analysis to identify specific compounds contributing to these alterations. We identified a total of 56 metabolites with an FDR-adjusted p-value of q ≤ 0.1 in at least one of the comparisons between cohorts (**Supplemental Data S3**). Notably, these alterations were not randomly distributed but predominantly affected specific metabolic classes, with a strong enrichment of lipid species. In particular, triacylglycerides were consistently upregulated in patients with long COVID compared to the control group of healthy survivors (**Figure 2A**). In addition to differences between long COVID patients and controls, we also observed metabolic distinctions between the two control groups (healthy survivors vs. uninfected individuals), highlighting that prior infection alone may have lasting metabolic effects (**Figure S3**). Moreover, several acylcarnitines, key intermediates of overarching mitochondrial fatty acid metabolism and energy production, were differentially regulated. For example, acetylcarnitine, the most abundant acylcarnitine in human plasma (11), was among the most strongly decreased metabolites in the serum of long COVID patients (**Figure 2A** and **S4A**). In addition, several other non-lipid metabolites with important physiological and pathological functions were differentially regulated, including homocysteine (Figure 2A and S5), dehydroepiandrosterone sulfate (**Figure 2A** and **S5**), several bile acids such as taurochenodeoxycholic acid (TCDCA) and taurocholic acid (TCA) (**Figure 2A** and **S5**), and proline betaine, a derivative of the amino acid proline (**Figure 2A** and **S5**). Importantly, these metabolite-level changes converged on biologically coherent, overarching pathways, as enrichment analysis of SMPDB (The Small Molecule Pathway Database) revealed significant enrichment of pathways related to homocysteine metabolism, acylcarnitine and fatty acid beta oxidation, and bile acid synthesis (**Figure 2B**).

**Figure 2.**
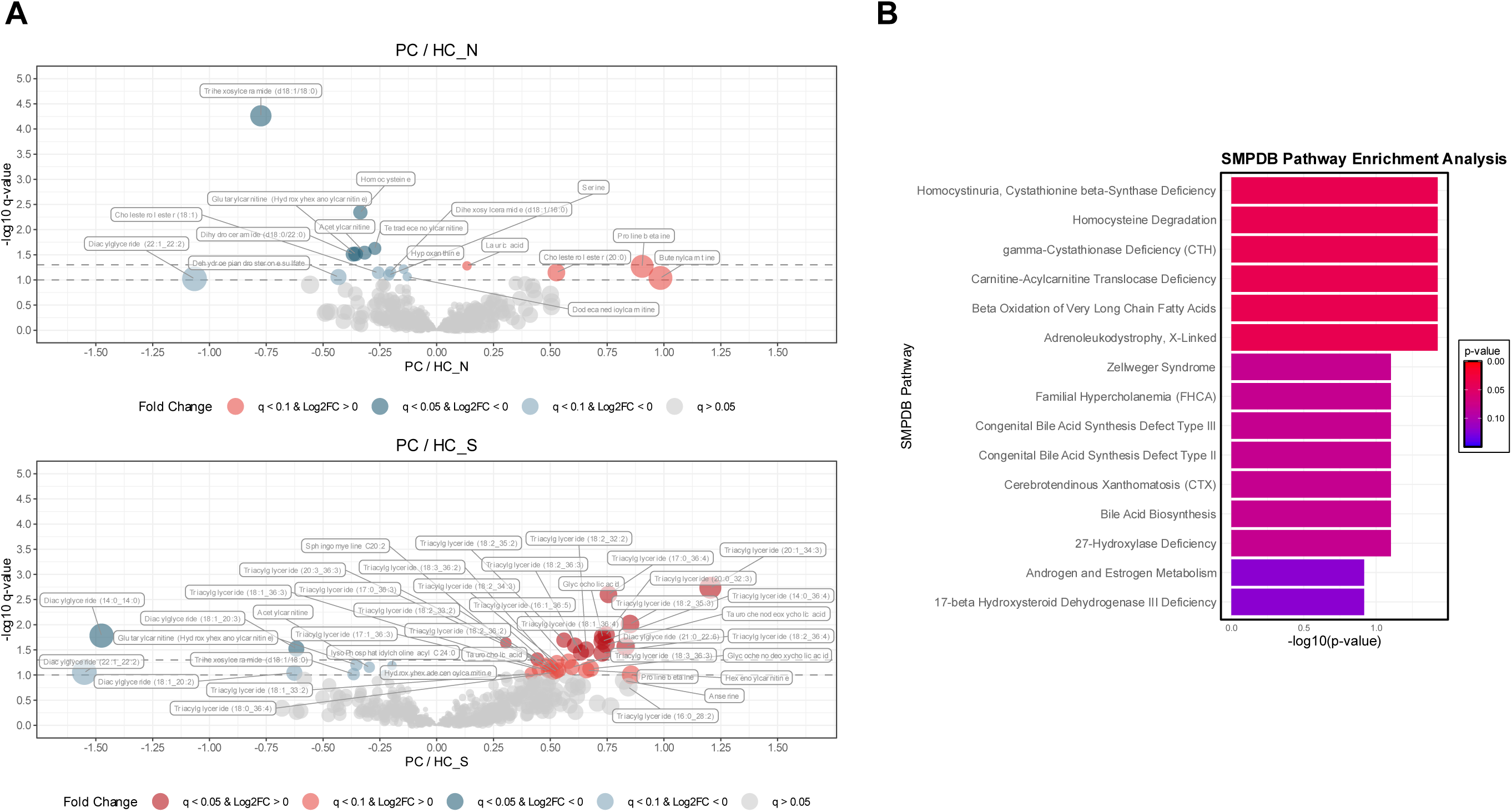
Volcano plots of the comparisons between long COVID patients and healthy control cohorts and pathway enrichment. **A** Volcano plots depicting the log_2_FC values (x-axis) and q-values (= FDR-adjusted p-values, y-axis) for all detected and quantified metabolites in the comparisons PC vs. HC_N (upper plot) and PC vs. HC_S (lower plot). The horizontal dashed lines indicate the different q-value cut-offs of q ≤ 0.1 and 0.05. Differentially abundant metabolites (DAMs) are coloured as indicated in the colour legend below the plot, all non-significant metabolites are coloured in grey. **B** Results of the SMDBP pathway enrichment analysis for the top 15 pathways. Shown are the negative decadic logarithms of the p-values from the enrichment results on the x-axis, on the y-axis are the enriched pathways depicted. In addition, the bars are coloured according to the p-values of the respective pathways, as indicated in the legend on the right side.

**Figure S3.**
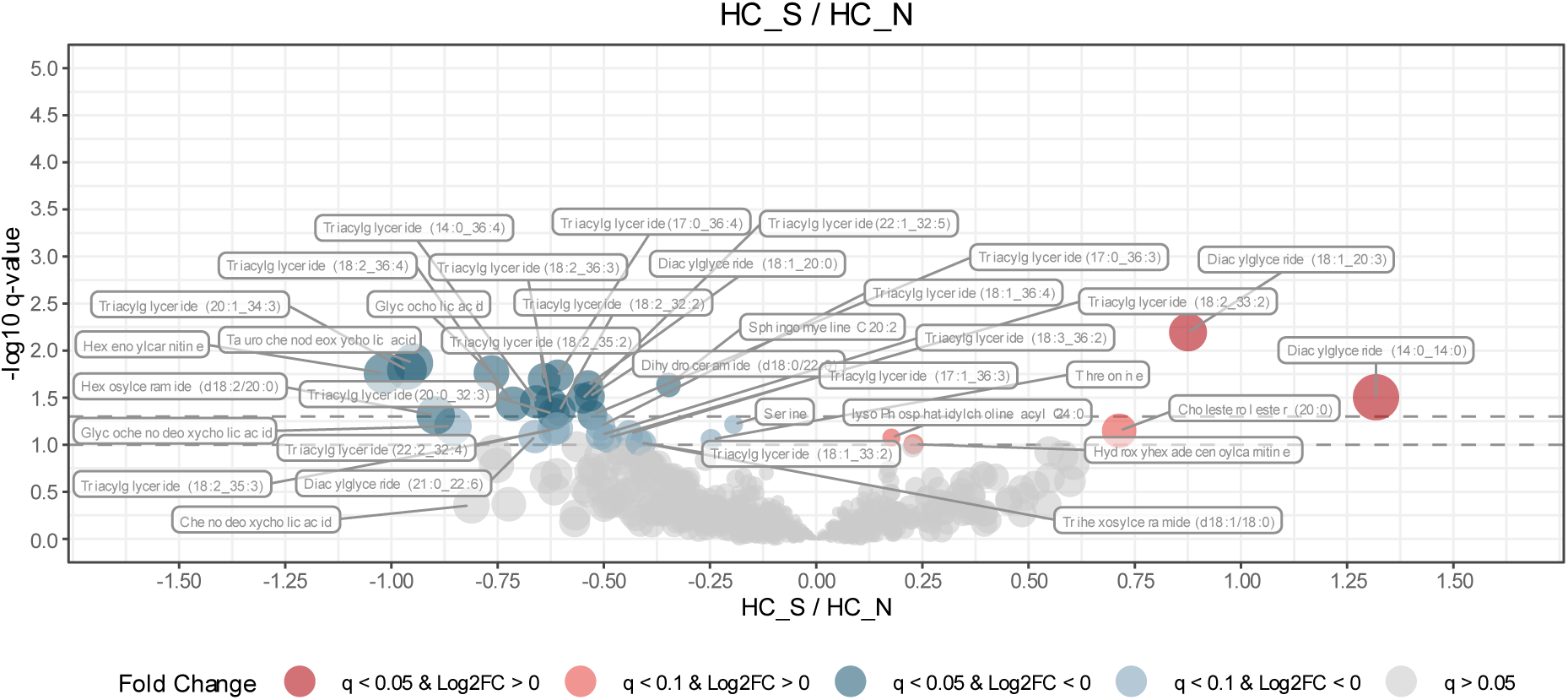
Volcano plot of the changes between HC_S and HC_N. **A** Volcano plot depicting the log2FC values (x-axis) and q-values (= FDR-adjusted p-values, y-axis) for all detected and quantified metabolites in the comparison HC_S vs. HC_N. The horizontal dashed lines indicate the different q-value cut-offs of q ≤ 0.1 and 0.05. Differentially abundant metabolites (DAMs) are coloured as indicated in the colour legend below the plot, all non-significant metabolites are coloured in grey.

### Association of differentially changed metabolites to neuropsychiatric symptom burden and cortical thickness in long COVID patients

Our results indicated PC-related differences in several lipid species and metabolites that are important for cellular energy homeostasis. Building on this, we aimed to investigate whether those PC-related metabolites were also associated with symptom severity in all cohorts, and particularly in the long COVID patients. Therefore, we next analyzed the association of the neuropsychological/psychometric scores obtained in this study (i.e. BDI-II, MADRS, MoCA, SF-36 GH and Vit) with variations in metabolite concentrations, while also taking cohort effects into account by including the interactions between the respective score and cohort effects. We determined a significant effect of each score by likelihood ratio tests of the model including the score by cohort interaction versus the model including cohort information only. After correcting for the false discovery rate, we found that no metabolites were significantly associated with changes in the BDI-II, MoCA or SF-36 GH, but in total 7 metabolites were associated with changes in the MADRS and 4 metabolites were associated with differences in the SF-36 vitality scores (**Figure 3**). Specifically, we identified one diacylglyceride (DAG (18:1_20:0)) and 6 more triacylglycerides (TAG (18:1_36:3), TAG (18:1_36:4), TAG (18:3_36:3), TAG (18:3_36:2), TAG (18:2_36:2) and TAG (18:2_36:3)) to be associated with changes in MADRS scores. Interestingly, while the overall estimate in the reference cohort (HC_N) was slightly negative (β = −0.06) for the overall fixed effect of the MADRS score on the DAG (18:1_20:0) levels, the effect of the MADRS score in the long COVID patients (PC) was slightly positive (β = 0.06), though both effects were not significant (p > 0.1). Moreover, for all triacylglycerides, the effects were the opposite, i.e. while the overall effect of the MADRS score on TAG levels was positive in the reference cohort, the effect was the opposite for the interaction between the MADRS and long COVID cohort (PC), e.g. for TAG (18:1_36:3) the overall fixed effect of MADRS score on metabolite concentrations in the reference cohort was β = 0.11 (p = 0.07) and in the long COVID cohort, i.e. the interaction effect of MADRS and cohort(PC) on TAG (18:1_36:3) levels was β = −0.13 (p = 0.04). These observations are also reflected in the inspection of the clustered z-score transformed concentrations, where hierarchical clustering was also performed within cohorts: In the long COVID cohort, individuals with higher TAG concentrations and lower MADRS scores cluster together (**Figure 3**). The opposite appears to be true in the healthy control groups, where individuals with higher TAG concentrations and also higher MADRS scores cluster together (**Figure 3**), although it should be noted that the MADRS scores in these healthy control cohorts were all below a clinically relevant threshold (**Figure S1**). In the analysis to the SF-36 vitality score as a measure for fatigue, we identified also one diacylglyceride (DAG (14:0_14:0)) to be associated with changes in SF-36 Vit. Specifically, there was a weak negative interaction between the overall SF-36 Vit score and DAG (14:0_14:0) concentrations in the reference cohort (β = −0.05, p = 0.01) with the opposite direction of this effect in the long COVID cohort (β = 0.04, p = 0.06). Hydroxyhexadecenoylcarnitine showed only a very weak association with SF-36 Vitality score (β = 0.01, p = 0.01), also the analysis for the two bile acids taurocholic acid (TCA) and taurochenodeoxycholic acid (TCDCA) showed only weak effects in the reference cohorts (β = −0.02, p = 0.04 for TCA and β = −0.03, p = 0.03 for TCDCA) (**Supplemental File S5** and **Figure 3**).

**Figure 3.**
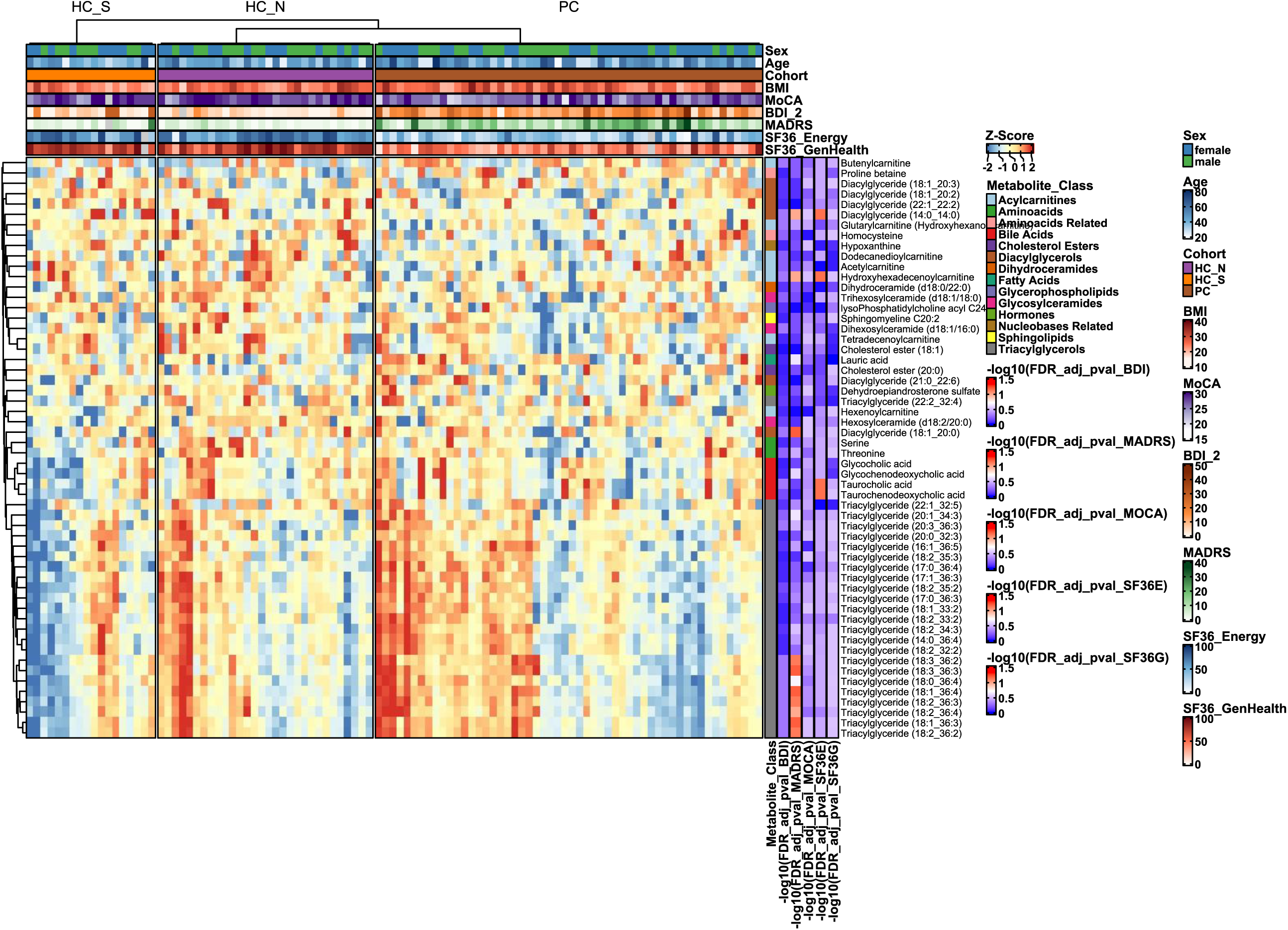
Heatmap with signatures of differentially changed metabolites and their association to neuropsychological/psychometric scores. **A** Visualization of the z-score normalized metabolite concentrations of all metabolites that were differentially regulated (q ≤ 0.1) in at least one of the three comparisons (HC_S/HC_N, PC/HC_N, PC/HC_S) of all samples after adjusting for batch, age, sex and BMI. Metabolites were clustered by agglomerative hierarchical clustering with average linkage. In addition, within each cohort, clustering was performed by default with agglomerative hierarchical clustering with complete linkage. In addition, several column and row annotations are shown in the heatmap: The columns are divided by cohort, the covariates age, sex and BMI are shown on different colour scales. Furthermore, the clinical symptom scores (BDI-II, MADRS, MoCA, SF36_Vitality and SF36_General) are annotated on the columns above each heatmap. Moreover, the negative decadic logarithms of the FDR-adjusted p-values of a χ2 difference test between two models with or without the respective symptom scores are shown. Lastly, the metabolite class of each metabolite is annotated to the right of the heatmaps.

Furthermore, we analyzed, in a methodically identical way to the analysis of the scores, the extent to which the cortical thickness of all regions of the Desikan-Killiany (DK40) atlas was associated with the blood level of the metabolites that significantly changed between the cohorts. Interestingly, we found that nearly all metabolites that were significantly associated with the MADRS or SF36-Vit score (**Figure 3**), showed also a significant link to different brain regions. Specifically, by hierarchical clustering of the −log_10_(q-values), we could identify one large cluster (“C1”) comprised of left- and right-hemispheric superiortemporal and middletemporal gyri and left-hemispheric insula in addition to right-hemispheric fusiform gyrus to be associated with blood metabolite concentrations of various triacylglycerides, but also diacylglycerides, hexosylceramide (d18:2/20:0) and acylcarnitines such as hexenoylcarnitine and tetradecenoylcarnitine (**Figure 4A**). Another acylcarnitine, hydroxyhexadecenoylcarnitine, was found to be associated with right hemispheric parahippocampal gyrus and the cuneus and hexenoylcarnitine showed also an association to the general mean thickness and inferiorparietal and superiorparietal gyri (**Figure 4A**). Furthermore, diacylglyceride (18:1_20:0), which was associated with changes in the MADRS score (**Figure 3**), was also found to be associated with cortical thickness in 4 regions: left hemispheric medialorbitofrontal and posteriorcingulate as right hemispheric isthmuscingulate and caudalmiddlefrontal gyri (**Figure 4A**). In addition, diacylglyceride (14:0_14:0), which was also found to be linked to differences in MADRS and SF36-Vit scores (**Figure 3**), showed an association to the cortical thickness with the left hemispheric superiorfrontal gyrus, right hemispheric insula and supramarginal gyrus (**Figure 4A**). To determine the direction of the identified associations, we also examined the estimates of the fixed effects for the respective DK40 regions in general and for the interactions between cortical thickness and cohort affiliation and plotted these estimates for all metabolites and regions for which at least one significant association was identified (p_adj(fdr)_ ≤ 0.1). For almost all metabolites significantly associated with C1, we found negative estimates for the overall effect of cortical thickness of the specific region on the respective metabolite in the reference cohort of healthy controls, except for hexosylceramide (d18:2/20:0) and hexenoylcarnitine, for which positive estimates were calculated. The regions that were only associated to hexenoylcarnitine serum concentrations, showed primarily positive estimates as well as for the regions associated with serum concentrations to diacylglyceride (18:1_20:0) and diacylglyceride (14:0_14:0) only negative estimates were calculated (**Figure 4B**). Strikingly, nearly all described estimates of the associations of the different regional cortical thicknesses to the aforementioned metabolites showed an opposite effect in the long COVID cohort, i.e. for the fixed effect of the interaction between the cortical thickness and cohort PC (**Figure 4C, Figure S4**). Moreover, apart from a few exceptions, this effect of reversed effect estimates could also be observed in comparison to the interaction between the cortical thickness and cohort HC_S (**Figure S5)**.

**Figure 4.**
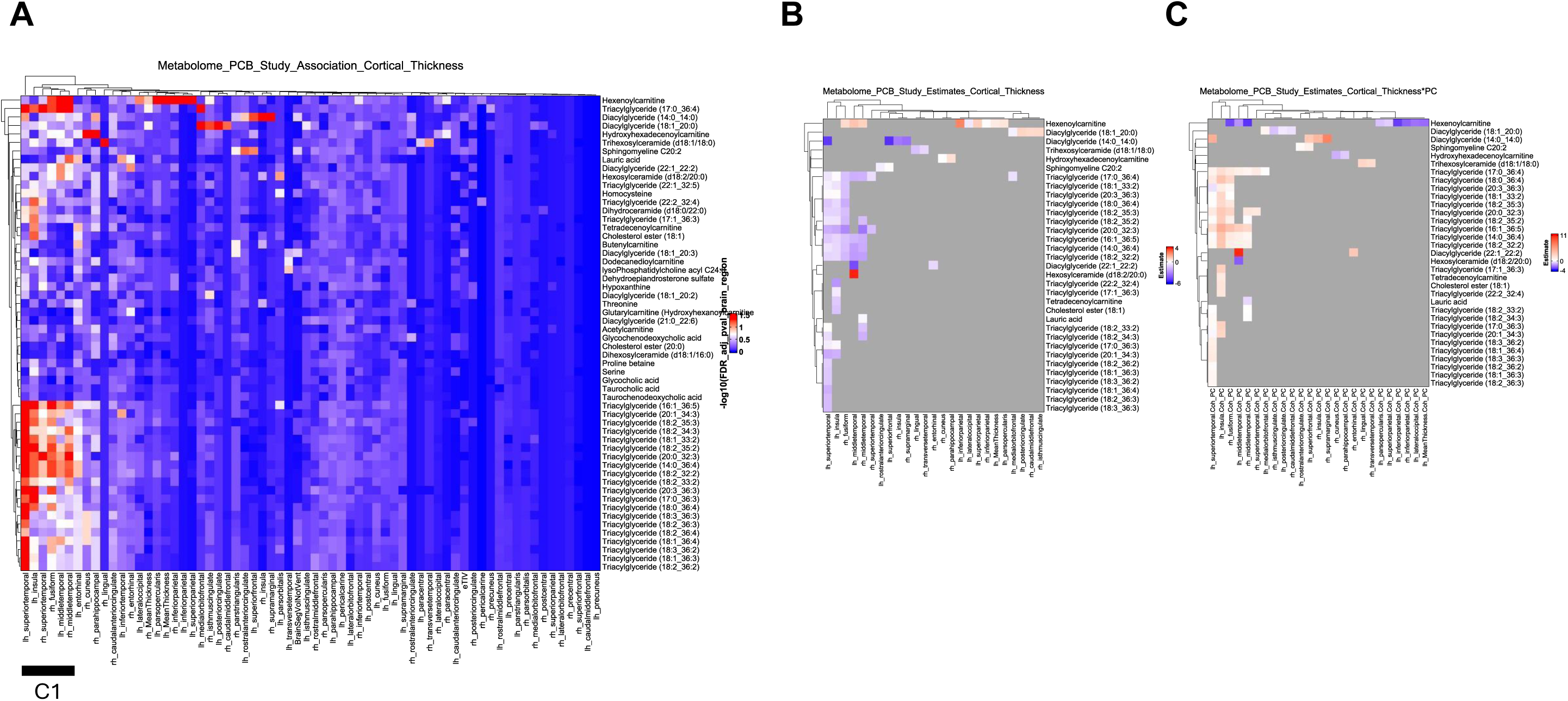
Heatmaps of FDR-adjusted p-values and estimates of metabolites significantly associated with cortical thickness. **A** Visualization of the negative decadic logarithms of the FDR-adjusted p-values of a χ2 difference test between two models with or without the cortical thickness of the respective brain region are shown, “lh” = left hemisphere, “rh” = right hemisphere. **B** and **C** Visualization of the estimates for the fixed effects of either only the cortical thickness across brain regions with changes in the concentration of the respective metabolite (**B**) or of the interaction between cortical thickness and cohort PC (**C**). The colour scales differ in **B** and **C** as they were dynamically computed according to the minimum and maximum estimate for the respective effect. Only the estimates of those metabolites for which the χ2 difference test yielded an FDR-adjusted p-value of ≤ 0.1 for association with the corresponding brain region are shown on a colour scale, all other estimates are excluded and are thus coloured grey.

**Figure S4.**
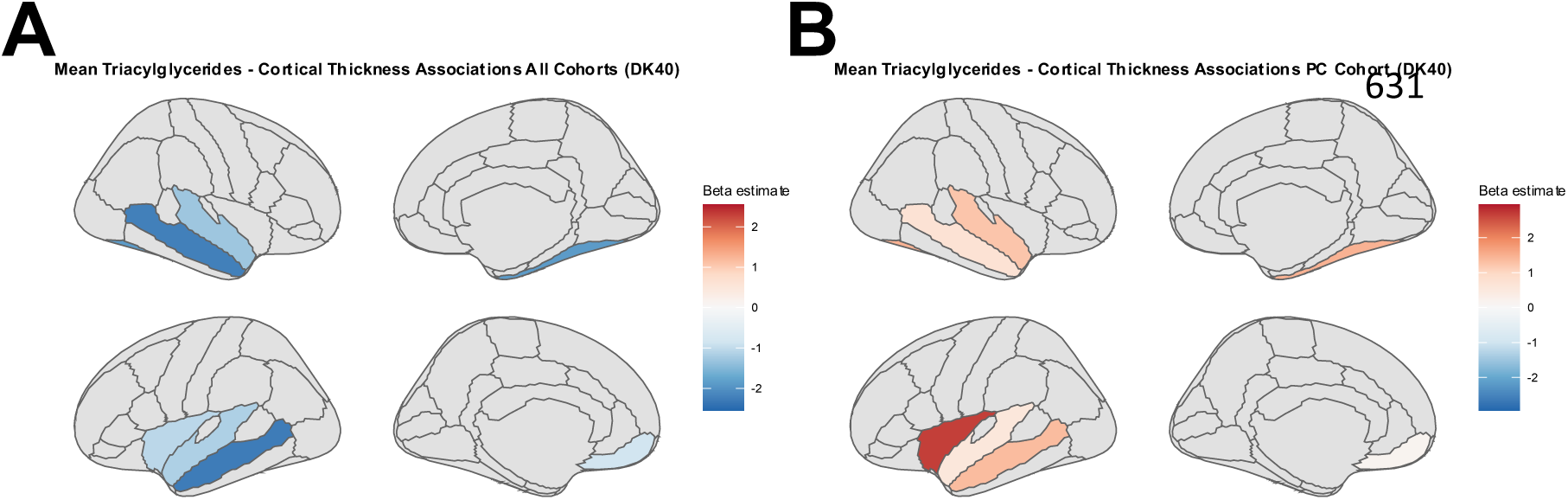
Mean estimates of significant associations between all triacylglycerides and cortical thickness. **A, B** Visualization of the mean estimates of all significant associations between all triacylglycerides and cortical thickness from DK40 atlas regions across all cohorts (**A**) or of the interaction between cortical thickness and cohort PC (**B**). Only the estimates of those metabolites for which the χ2 difference test yielded an FDR-adjusted p-value of ≤ 0.1 for association with the corresponding brain region are shown on a colour scale, all other estimates are excluded and respective regions are thus coloured grey.

**Figure S5.**
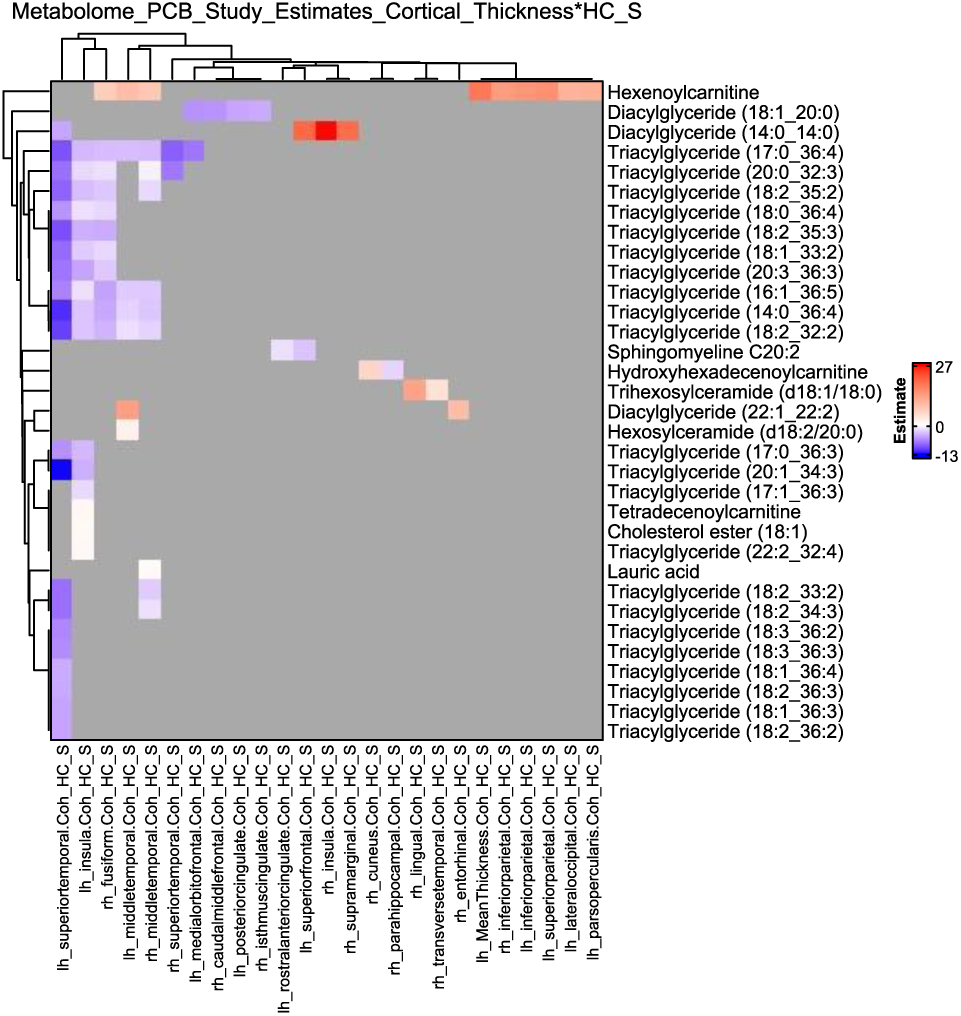
Heatmaps of estimates of the interaction between the cortical thickness and cohort HC_S of metabolites significantly associated with cortical thickness. Visualization of the estimates for the fixed effects of the interaction between cortical thickness and cohort HC_S across brain regions (“lh” = left hemisphere, “rh” = right hemisphere) with changes in the concentration of the respective metabolite. Only the estimates of those metabolites for which the χ2 difference test yielded an FDR-adjusted p-value of ≤ 0.1 for association with the corresponding brain region are shown on a colour scale, all other estimates are excluded and are thus coloured grey.

### Validation of the longCOVID peripheral metabolite signature in an independent cohort

We wanted to determine whether our observations could be replicated in an independent cohort. Therefore, we took the - to our knowledge - largest metabolomics dataset from Su et al. (15) and examined alterations in the differentially altered metabolites identified in our study in long COVID patients in their INCOV cohort of patients with PASC (≥ 1 PASC symptom according to the clinical information from Table S1, sheet “S1.3 PASC data” (15)). Of all 56 metabolites that were differentially regulated in our study, 13 metabolites were identified, that were also measured in the Su et al. study. Interestingly, most of the metabolites (10 out of 13) that we identified as being different in long COVID patients showed a similar pattern of regulation in their study and, moreover, for 9 out of 13 in the same direction (i.e. up- or downregulation) as in our cohort (**Figure 5**, threonine was not altered in the Su et al. dataset nor in the comparisons of PC with HC_N or HC_S in our dataset). Different results were identified for hypoxanthine (upregulation in Su et al. dataset, downregulation in the comparison PC vs. HC_N in our study), lauric acid (downregulation in Su et al. dataset, upregulation in the comparison PC vs. HC_N in our study) and serine (upregulation in Su et al. dataset, downregulation in the comparison PC vs. HC_N in our study).

**Figure 5.**
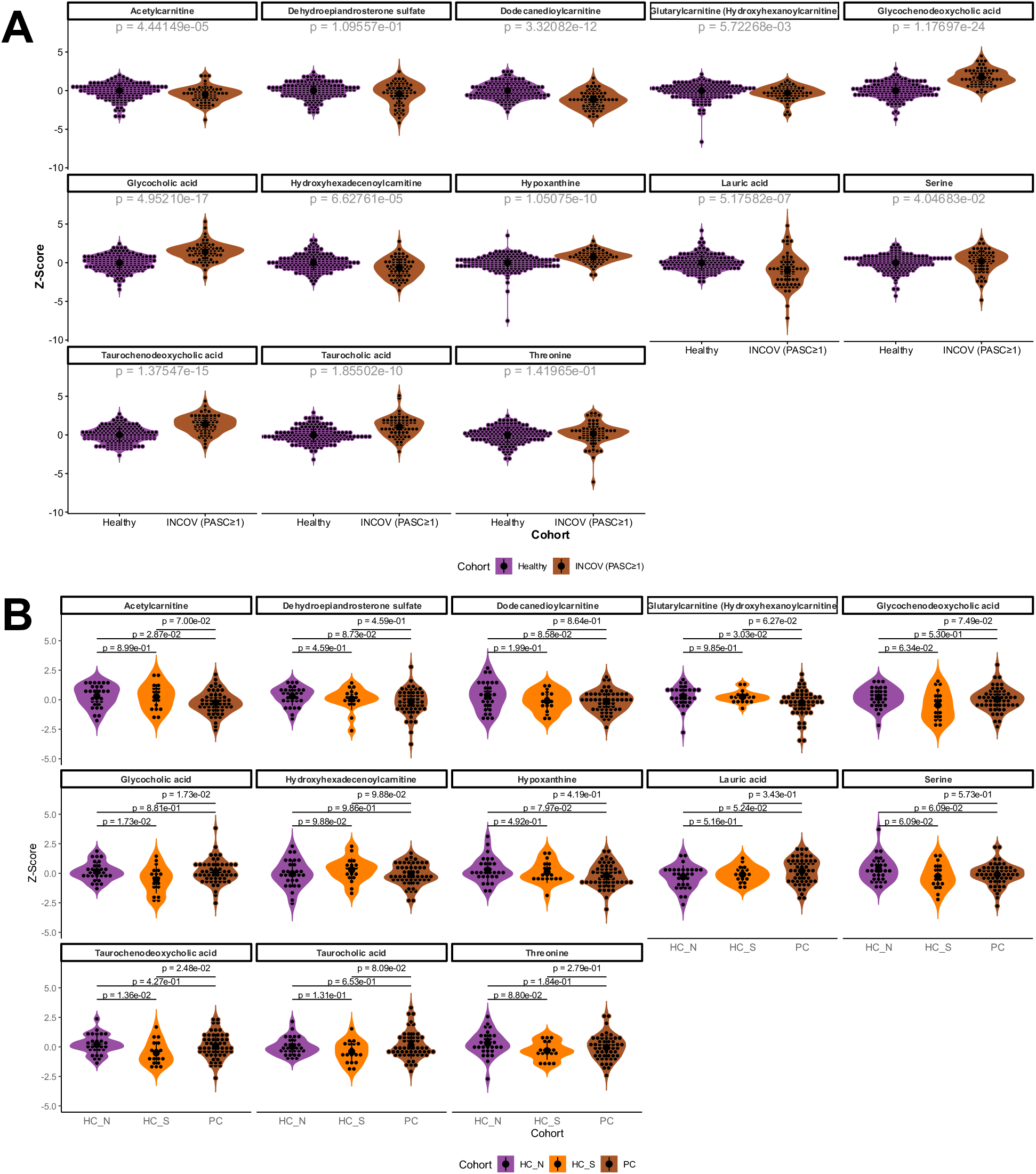
Validation of differentially changed metabolites in an independent cohort. **A** Z-score transformed metabolite concentrations, adjusted for age, sex and BMI, were retrieved from Su et al. (15) for all detected metabolites, that were significantly (q ≤ 0.1) regulated in at least one of the three comparisons (HC_S/HC_N, PC/HC_N, PC/HC_S) in our cohort. The cohorts were divided in the healthy controls (n = 178) and all patients from the INCOV cohort with ≥ 1 PASC symptom (n = 68). The shown metabolomic data are from time point T3, i.e. 2-3 months after initial infection. The shown p-values represent the FDR-corrected exact p-values from two-tailed Mann-Whitney tests. **B** Violin plots showing batch and covariate (age, sex and BMI) adjusted and z-score transformed concentrations of all metabolites from this study that were significantly changed (q ≤ 0.1) between the different cohorts and also detected by Su et al. (15). The q-values represent the FDR-adjusted p-values of post-hoc Tukey tests.

## IV. Discussion

In this study, we used targeted metabolomics to identify biological factors that are altered in long COVID patients and that may shed light on potential underlying pathophysiological processes of PASC. The cohort we studied was well phenotyped and showed a mild depressive phenotype, mild cognitive deficits and increased fatigue. We revealed subtle, but biologically coherent alterations, with increases in tri-and diacylglycerides as compared to healthy survivors and a decrease in several acylcarnitines, including acetylcarnitine. Circulating tri- and diacylglycerides serve as an energy source for all tissues, as they are taken into cells after hydrolysis by lipoprotein lipase (LPL) (42). An increase in the blood stream may thus reflect a lower uptake into cells due to decreased LPL activity, a well-known mechanism in patients with insulin resistance (43). In fact, insulin resistance and decreased LPL activity can be driven by chronic inflammation through elevated circulating cytokines, such as in patients with obesity by interferon-γ (44), which is one of the most important antiviral cytokines induced during viral infections (45). IFN-γ has also been identified to be of high importance for the host response to an acute Sars-CoV-2 infection (46) and has been identified to be not only persistently increased also in long COVID patients, but also driving inflammatory serum signatures in a subset of long COVID patients with especially fatigue as PASC symptom (47) and was upregulated in long COVID patients with cognitive deficits (26). Interestingly, an upregulation of triacylglycerides has also been observed in a large number of patients with ME/CFS in the UK Biobank, where ME/CFS patients showed the highest TAG levels in total, followed by depressive patients and patients with hypertension (48). In addition to the upregulation of TAGs in depressive patients, obesity itself, i.e. a higher BMI, has previously been shown to be associated with a more chronic course of the disease in MDD patients (49) and also with a reduction in temporo-frontal cortical thickness (50). We also found in this study that levels of TAGs that were – after controlling for BMI – upregulated in long COVID patients, were not only associated to primarily associated with cortical thickness in temporal regions (**Figure 4**). It should be noted, that this association was positive in the cohort of long COVID patients, i.e. increased levels of those TAGs that were upregulated in the long COVID patient group corresponded to a bigger cortical thickness in regions that were found to show larger cortical thickness due to potentially ongoing inflammation and tissue swelling in long COVID patients in a previous study (26). However, the interaction effect between long COVID cohort and MADRS scores as indicator of depressive symptoms on TAG levels was negative, which contrasts with most large-scale studies that showed positive associations between higher TAG levels and depression (51) and highlights on the one hand one limitation of our study, a relatively small sample size, and on the other hand the need for larger studies investigating potential associations between lipid species and depressive symptoms in long COVID patients. Another important class of molecules that was differentially regulated in long COVID patients compared to healthy control cohorts were acylcarnitines, which are important regulators of fatty acid import into mitochondria and thus beta-oxidation (11). We have already identified a downregulation of acetylcarnitine as a short-chain acylcarnitine in the data set of Su et al. (15) in a previous manuscript (16) and could also identify this downregulation in our cohort of long COVID patients. Acetylcarnitine has been found to be also downregulated in multiple other neuropsychiatric diseases, including MDD and ME/CFS and it’s supplementation led to reduced fatigue and depressive symptoms in several clinical trials in patients with ME/CFS and MDD (16). This observation is of interest in light of the identified upregulation of TAGs in long COVID patients, as lower levels of acetylcarnitine might reflect a decrease in beta-oxidation which might contribute to the observed higher levels of TAGs and also decreased mitochondrial energy productions as it can be frequently observed in ME/CFS patients with fatigue (9). Furthermore, we identified also hexenoylcarnitine and butenylcarnitine to be upregulated in long COVID patients, which are unsaturated medium-chain acylcarnitines (11), that have been found to be upregulated in individuals with obesity (52, 53) and may indicate disturbance of branched-chain amino acid metabolism or may be related to altered production of microbial metabolites (11). While it was upregulated in long COVID patients, we found a negative association between cortical thickness and hexenoylcarnitine levels in those patients for several tempo-parietal regions. However, while no reports exist to our knowledge about any association of hexenoylcarnitine, butenylcarnitinie and other branched-chain acylcarnitines to structural brain changes, severe accumulation of branched-chain acylcarnitines leads in newborns with short/branched-chain acyl-CoA dehydrogenase (SBCAD) deficiency to seizures and developmental delay. In addition to the identified perturbations in lipid metabolism, we also found multiple other metabolites to be altered in long COVID syndrome, such as several bile acids, e.g. taurochenodeoxycholic acid (TCDCA), taurocholic acid (TCA), glycochenodeoxycholic acid (GCDCA) and glycocholic acid (GCA) were found to be upregulated as compared to the control cohort of healthy survivors (**Figure 2** and **5B**), all of which were also upregulated in the INCOV cohort of PASC patients (**Figure 5A**) (15) and TCA and TCDCA were also weakly associated with fatigue (SF36-Vit, **Figure 3**). All of these are primary bile acids that are primarily produced in the liver and circulate after reabsorption in the blood stream and can enter peripheral tissues, such as the CNS via passive diffusion and transport proteins (54). All said bile acids bind and activate the sphingosine-1-phosphate receptor-2 (S1P2R), which can induce microglia activation and neuroinflammation as it has been shown in a mouse model of hepatic encephalopathy, in this case specifically for TCA (55). Moreover, as recently reviewed and summarized by Lirong et al. (56), the bile acids we identified to be upregulated in the blood of long COVID patients in our dataset and in the one from Su et al. (15), show also higher blood levels in stroke patients (GCDCA) (57), Alzheimer’s disease (GCA, GCDCA and TCA, TCDCA) (56, 58, 59), as well as an increased ratio of GCDCA to CA (cholic acid) in post-mortem brain samples has been demonstrated recently (59), indicating also an increased presence and induction of potentially neuroinflammation. In addition, GCA was also upregulated in a mouse model of depression (56) and correlated positively anxiety and depressive symptom severity in patients with Crohn’s disease (60). It should be noted that patients with cholestasis such as in primary biliary cholangitis (PBC) often exhibit fatigue as a symptom (61) and bile acids such as TCA, TCDCA, GCA and GCDCA are commonly found upregulated in PBC patients (62) though – to our knowledge – no studies exist that have proven a causal relationship between bile acid upregulation and fatigue severity. Taken together, our results show primarily differential abundance in metabolites that may indicate potential disturbances in fatty acid metabolism and subsequently mitochondrial energy production – as it has been shown in ME/CFS patients (9, 12) – that might also correspond to changes in cortical thickness in long COVID patients, that were described in detail in a previous study from our group (26).

This study also has several limitations. First of all, an important limitation is the small sample size, which may impair – given the potentially considerable heterogeneity of biological subtypes of long COVID syndrome (47, 63, 64) – the ability to detect both, between-cohort differences as well as associations to other biological variables. This is important to bear in mind when interpreting our results, because we also found differences between our control groups, i.e. between the healthy survivors and the healthy uninfected individuals, which need to be investigated in further studies with larger cohorts. In addition, compared to the study by Su et al. for example, our study cohort not only consisted of patients with PASC 2 to 3 months after initial infection, but the patients also showed a greater heterogeneity in terms of duration of PASC symptoms, which may not only influence the concentration of metabolites, which vary up to 2 years after initial infection (65) but may also explain some of the different results of our analyses compared to the Su et al. metabolome (15), though the majority of metabolites detected in both studies showed similar profiles in the PASC/long COVID cohort (**Figure 5**). In this study, we used a targeted metabolomics approach and analysed a pre-defined panel of metabolites, which obviously introduces a strong selection bias, as a major part of this panel is a large set of lipids, which naturally increases the probability of detecting differences in lipid concentrations. Therefore, we would like to emphasise that the results obtained should be treated with caution and need to be extended by larger, unbiased studies with larger cohort sizes and untargeted mass spectrometry-based metabolomics. Lastly, all our results do allow only speculations about whether the determined metabolome alterations in long COVID patients may causally contribute to the disease phenotype or are rather only the consequence or compensation of other systemic perturbations.

Nevertheless, our results uncover metabolomic alterations in long COVID patients and provide for the first time an association of some of these metabolomic alterations in depressive, cognitive and fatigue symptom severity as well as cortical thickness in important brain regions important for affective and cognitive symptoms (26). These findings unveil further somatic correlates of neuropsychiatric PASC symptoms and may extend the biological knowledge of long COVID syndrome.

## V. Supplementary tables

**Supplemental Dataset S1 Baseline demographics and neuropsychological/psychometric scores for all study participants**

**Supplemental Dataset S2 Batch-corrected, z-score transformed metabolomics data, adjusted for age, sex and BMI**

**Supplemental Dataset S3 Log_2_ Fold Changes and FDR-adjusted p-values for all metabolites from all between-cohort comparisons**

## Acknowledgments

We are grateful to Ines Krumbein for overseeing MRI measurements and to Lara Krickow, Maximilian Vollmer and Marlene Müller for collection of MRI data and pre-analytics of blood samples. Open access funding provided by Projekt DEAL.

## Conflict of Interest

MW is a member of the advisory boards and gave presentations for the following companies: HMNC, Janssen Pharmaceutical Research, Novartis, Boehringer Ingelheim, Germany; Bayer AG, Germany; and Biologische Heilmittel Heel GmbH, Germany. MW has further conducted studies with institutional research support from HEEL and from Janssen Pharmaceutical Research for a clinical trial (IIT) on ketamine in patients with major depression unrelated to this investigation. BB received industrial funding for a different Post-COVID study from AlzChem Group (Trostberg, Germany) unrelated to this analysis.

## Funding

The project on which this report is based was funded by the Federal Ministry of Education and Research under grant number 01EQ2403A (to MW). The responsibility for the content of this publication lies with the author. In addition, this project was also funded and by a Clinician Scientist Grant to DLB (IZKF, CSP032) and by an Advanced Clinician Scientist Grant to BB (IZKF, ASCP001).

## Data availability

The datasets generated and/or analyzed during the current study are available from the corresponding and the first author on reasonable request.

## Notes

### Author Declarations

All participants provided written informed consent before participating in the study. The study protocol was approved by the local Ethics Committee of Jena University Medical School.

